# Characteristics of Suicide Prevention Apps: A Content Analysis of Apps Available in Canada and the United Kingdom

**DOI:** 10.1101/2024.07.10.24310091

**Authors:** Laura Bennett-Poynter, Samantha Groves, Jessica Kemp, Hwayeon Danielle Shin, Lydia Sequeira, Karen Lascelles, Gillian Strudwick

**Author notes:** Corresponding Author: Dr. Gillian Strudwick RN, PhD, FAMIA, FCAN, Campbell Family Mental Health Research Institute, Centre for Addiction and Mental Health 60 White Squirrel Way, Toronto, Ontario, Canada. Denotes joint first authors (Laura Bennett-Poynter, Samantha Groves).

## Abstract

**Objective:** We aimed to examine the characteristics, features, and content of suicide prevention mobile apps available in app stores in Canada and the United Kingdom.

**Design:** Suicide prevention apps were identified from Apple and Android app stores between March-April 2023. Apps were screened against predefined inclusion criteria, and duplicate apps were removed. Data were then extracted based on descriptive (e.g., genre, app developer), security (e.g., password protection), and design features (e.g., personalization options). Content of apps were assessed using the Essential Features Framework. Extracted data were analyzed using a content analysis approach including narrative frequencies and descriptive statistics.

**Results:** Fifty-two (n=52) suicide prevention apps were included within the review. Most were tailored for the general population and were in English language only. One app had the option to increase app accessibility by offering content presented using sign language. Many apps allowed some form of personalization by adding text content, however most did not facilitate further customization such as the ability to upload photo and audio content. All identified apps included content from at least one of the domains of the Essential Features Framework. The most commonly included domains were sources of suicide prevention support, and information about suicide. The domain least frequently included was screening tools followed by wellness content. No identified apps had the ability to be linked to patient medical records.

**Conclusions:** The findings of this research present implications for the development of future suicide prevention apps. Development of a co-produced suicide prevention app which is accessible, allows for personalization, and can be integrated into clinical care may present an opportunity to enhance suicide prevention support for individuals experiencing suicidal thoughts and behaviours.

**Article Summary:** *Strengths and limitations of this study:*

- This app review used an established method for systematically identifying and examining suicide prevention apps, which has been successfully used previously.
- There is potential for overlap between different domains of the Essential Features Framework, which could lead to changes in reporting of percentages relating to app review findings.
- Only apps available in the UK and Canada in the English language were assessed. Current provision and content of suicide prevention apps may differ across countries, including those available in lower- and middle-income countries. Due to resource and time constraints, the quality of apps were not assessed.

## Introduction

Suicide is a global public health problem, where over 700,000 people die by suicide each year [1]. Suicide and self-harm are disproportionately prevalent among certain groups of individuals, including but not limited to individuals experiencing mental health conditions [2], middle aged men [3], adolescents [4], and LGBTQIA+ populations [5, 6].

Despite the high demand for suicide prevention resources, many individuals with suicidal thoughts or behaviours face barriers to receiving care. Individuals attempting to access primary or secondary mental health services may experience long waiting lists for treatment, inconsistent intervention provision across regions, and perceived or actual stigma associated with accessing care [7]. Additionally, individuals presenting to emergency departments following self-harm or a suicide attempt may face further barriers to appropriate care including a lack of time and space for a full psychosocial assessment, and inadequate staff training in mental health care [8]. Finally, worldwide, healthcare providers are experiencing staffing shortages, with projections of a potential 18 million shortfall of health workers by 2030, with low and middle-income countries being the most affected [9]. As a result of these issues, there is a need to consider modes to supplement suicide prevention care.

Mobile apps may offer a novel adjunct to suicide prevention interventions delivered by health, social care, or voluntary service practitioners. Mobile device use is common in the general population. As of 2022, estimates suggest 86% of people in the UK, and 84% of people in Canada own a smartphone, with levels expected to increase in the coming years [10]. App based resources may assist in mitigating the detrimental impacts of staffing shortages, waitlists for treatment, and social distancing measures [11, 12]. Furthermore, benefits for users may include anonymity, cost-effectiveness, and the ability to use interventions at the user’s own pace [13]. A meta-analysis analyzing the efficacy of self-guided suicide prevention digital interventions (n=14) found that such web or app interventions had a small but significant effect on reducing suicide ideation, as long as apps were directly targeted to prevent suicide rather than depression generally [13].

Despite the promise of suicide prevention apps, previous studies have identified some issues with the apps currently available. Reviews of apps available in the United States and Australia have found a lack of adherence to clinical guidelines regarding provision of evidence-based support. For example, apps have been found to lack content such as psychoeducation, safety plans, and access to support networks or emergency/crisis support [14-16]. Furthermore, app availability has been shown to be unpredictable, with apps regularly becoming unavailable, and search results for relevant apps being highly variable upon date of searching [17]. In order to develop a novel suicide prevention app, there is a need to understand what is currently available to users, in this case for an app tailored for a UK and Canadian audience.

In recent years efforts have been made to design tools to allow systematic assessment of suicide prevention apps. One such tool is the Essential Features Framework [18] which aims to facilitate assessment of apps based on content, features, and design. This framework was developed using a systematic review of articles examining development, implementation, feasibility, or effectiveness of suicide prevention apps, where features of apps discussed were synthesized using thematic analysis. The framework is comprised of eight domains 1) General information regarding suicide; 2) Wellness; 3) Positivity and inspiration; 4) Distraction and alternate activities; 5) Safety planning; 6) Screening tools; 7) Helpful resources; and 8) Immediate help-seeking. Definitions of each framework domain and examples of eligible features are presented in Table 1. No published studies have yet used this framework to assess the content of suicide prevention apps.

**Table 1.** Essential Features Framework [18].

|  |  |
| --- | --- |
|  | Definition and Example Features |
| **General Information about Suicide** |  |
|  | Information about suicide including risk factors and warning signs, dispelling myths about suicide, and information regarding how to support someone who is suicidal. |
| **Wellness** |  |
|  | Tools to promote wellbeing such as mindfulness and relaxation exercises. |
| **Positivity and Inspiration** |  |
|  | Content such as inspirational messages, quotes from individuals with lived experience, and suggested reasons for living. |
| **Distraction and Alternate Activities** |  |
|  | Tools and suggestions to support users such as coping strategies and distraction activities. |
| **Safety Planning** |  |
|  | A tool to help someone cope with suicidal thoughts, or to support someone who experiences suicidal thoughts. Includes ways to recognize when someone may be nearing a crisis, coping strategies to use, and useful sources of support to contact (e.g., family, friends, clinicians, helplines) [20]. |
| **Screening Tools** |  |
|  | Tools used for identifying and monitoring mood, screening for psychological distress or “suicide risk”. |
| **Helpful Resources (for Help-Seeking)** |  |
|  | Information and contact details for suicide prevention helplines, mental health services, and emergency departments. |
| **Immediate Help-Seeking** |  |
|  | Fast access to immediate help such as links to suicide helplines and emergency services on the app homepage. |

## Objectives

We aimed to use the Essential Features Framework to review the characteristics and content of suicide prevention apps available within the UK and Canada to inform the development of an evidence-based suicide prevention app to be made available in these countries.

## Methods

### App Store Search

Development of search terms followed previous research [14, 19]. The search terms used were “*Suicide Prevention*”, “*Suicide*” and “*Safety Plan*” and no restrictions were imposed related to store subcategories. Searches of the ‘Apple App Store’ and ‘Google Play’ in the UK and Canada were conducted from 3rd March - 12th April 2023. The UK app search was conducted by SG (using a Google Pixel 6a) and LBP (using an iPhone 12 mini, iOS version 16.1.1). HDS (using an iPhone 11 Pro, iOS 15.6.1) and JK (using an iPhone 11, iOS version 16.4.1) conducted the Canadian app search.

### Selection Criteria

Identified apps were eligible for inclusion if they were: a) free, b) developed in the English language, c) could be downloaded on an Apple or Android device in England or Canada, d) the focus of the app was suicide prevention, and e) the target users of the app were individuals experiencing suicide-related thoughts and/or behaviours. General mental health apps or apps developed for a specific mental health condition were included if the app contained a suicide prevention resource such as a suicide safety plan. Similarly, apps tailored for individuals supporting someone with suicidal thoughts were included only if they had suicide prevention resources such as a safety plan, or the ability to call for emergency support for someone experiencing suicidal thoughts themselves. We excluded apps solely designed for healthcare providers.

Apps were initially screened independently for eligibility based on the app title and description visible on the app store page (LBP, SG, JK, HDS). A subsequent round of screening was conducted where apps were downloaded, and full content of apps screened against the eligibility criteria (LBP, SG, JK). Duplicate apps (e.g., if an app was identified by both UK and Canadian searches) were manually removed.

### Data Extraction

SG, LBP, and JK extracted data. A template data extraction form is shown in Supplementary File 1. Briefly, basic descriptive data were extracted (e.g., name, genre, developer of the app), alongside security (e.g., whether password protection available), and design (e.g., whether the app could be personalized) features. App content was examined using the Essential Features Framework [18]. Data extraction was performed by one researcher (specific person dependent on app location and whether the was available on Apple or Android) and was then verified by a second reviewer. Any conflicts in extraction were discussed including consultation with other members of the research team until consensus was reached. Quality of apps was not assessed.

### Analysis

A content analysis approach was conducted. This method has been previously used to evaluate suicide prevention apps [16]. A coding framework was developed using the Essential Features Framework, alongside content of preliminary discussions held during co-production workshops regarding perceived important features of suicide prevention apps. These workshops gained the perspectives of service users, informal carers, and clinicians. Extracted data were synthesized using narrative descriptions of app features, alongside frequencies and percentages of app characteristics (data analysis performed by LBP and SG). Ethical approval was not required.

## Results

### Search Results

Following de-duplication 716 apps were identified by searches. After title and description screening, 77 apps remained, and following full app assessment, 52 apps were included within the review (see Figure 1 for the PRISMA diagram for screening process and Supplementary File 2 for a list of included and excluded apps alongside reasons for exclusion). Search results were highly variable upon day of searching, with a further number of apps (n=14) becoming no longer available during data extraction. During extraction, the authors encountered malfunctions and errors in multiple apps (20, 38.5%), ranging from apps freezing during use, to erroneous links to outside resources and helplines.

**Figure 1.**
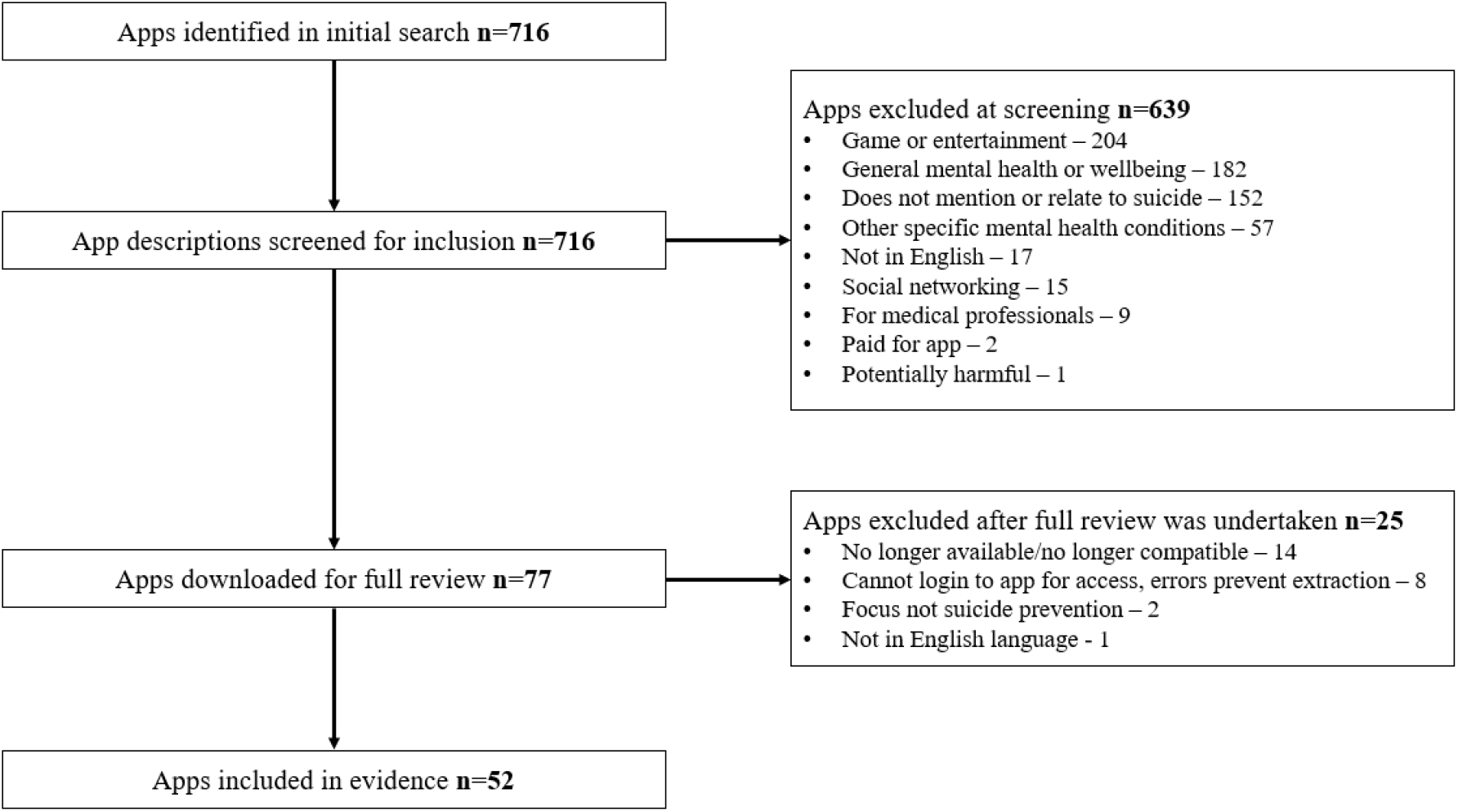
PRISMA diagram of app screening, review, and inclusion in evidence

### Basic App Characteristic

Included apps were mostly developed in the USA (32/52, 61.5%), with apps also developed in Australia (7/52, 13.5%), Canada (6/52, 11.5%), the UK (6/52, 11.5%), and Zimbabwe (1/52, 1.9%). Most were available in English only (45/52, 86.5%), and only one app had an accessibility feature (1/52, 1.9%), in this case by including content presented in British Sign Language. User ratings were available for some apps (15/52, 28.8%) with an average rating of 4.3 out of 5.0, among apps showing ratings. Data were available on Android apps regarding the number of downloads (N=41/52, 78.8%). The two most popular apps had been downloaded over 100,000 times. Android apps also stated the date of the most recent app update. Some apps (16/41, 39.0%) had not been updated in over 3 years. See Table 2 for basic characteristics of included apps.

**Table 2.**
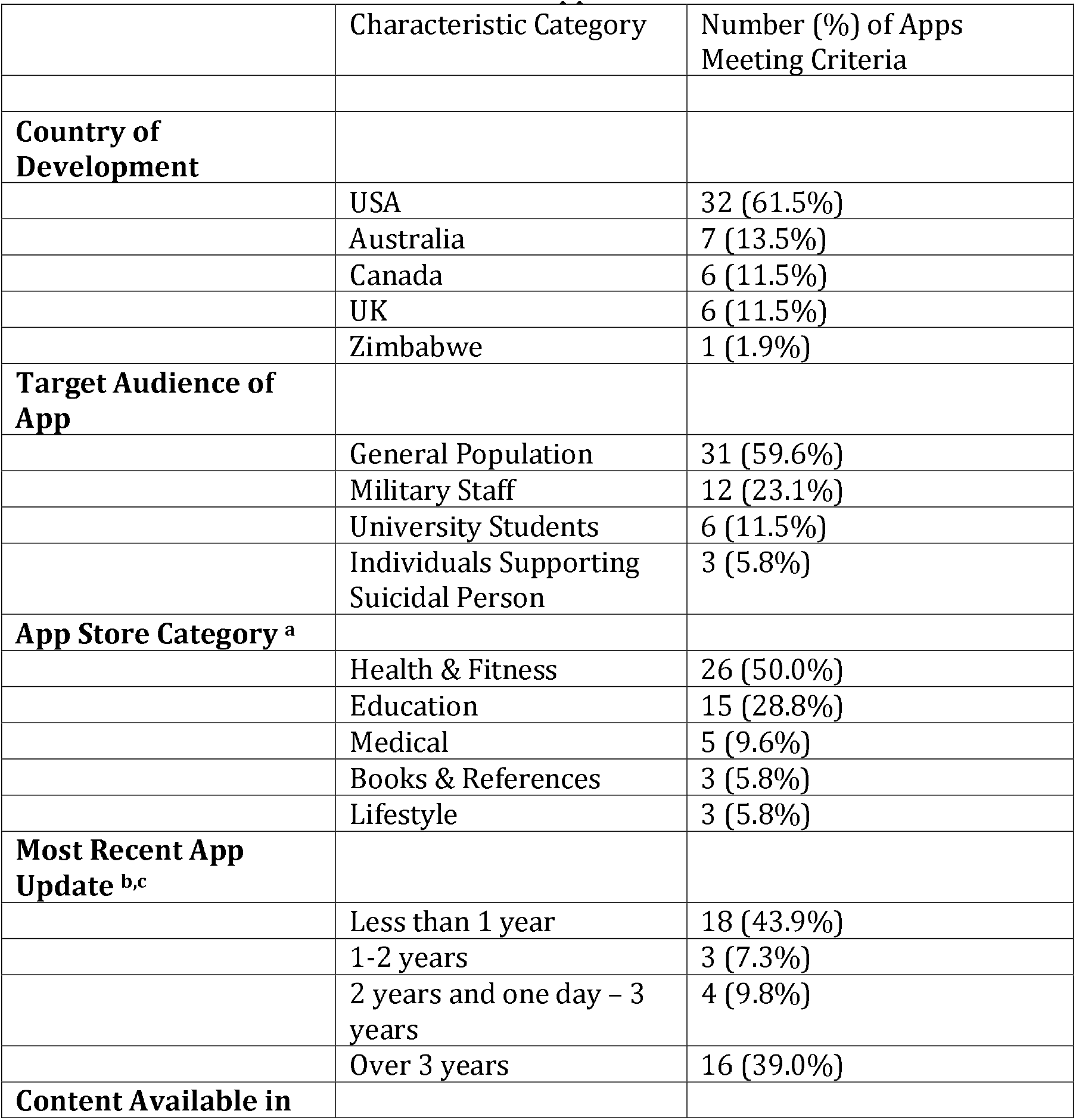
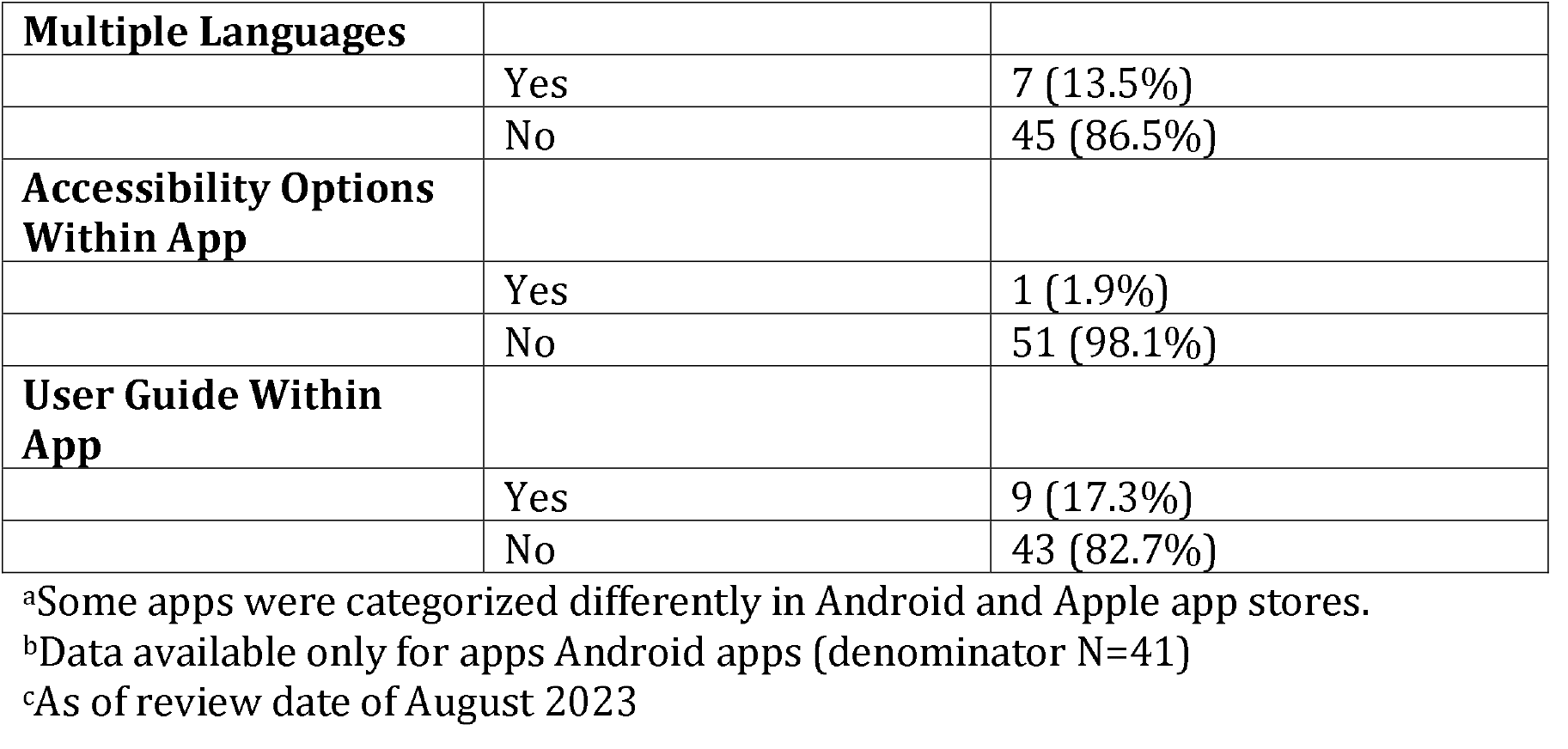
Basic Characteristics of Included Apps.

The majority of apps were tailored for the general population (31/52, 59.6%), with some of these specifically designed for young people (4/31, 12.9%). Additional apps were tailored to staff at military bases or veterans (12/52, 23.1%), students at specific universities (6/52, 11.5%), and individuals who are supporting, or are concerned about someone who may be suicidal (3/52, 5.8%). Despite the majority of apps being tailored for users experiencing suicidal thoughts (by necessity of inclusion criteria), almost three quarters of apps also contained content for those concerned about a person who may be suicidal (38/52, 73.1%). For example, some apps provided advice on what to say or avoid saying if someone shares that they are thinking about suicide, potential warning signs that a person may be suicidal, and suggestions of potential methods of support. Although some apps gave the user space to enter the contact details of clinicians for future reference, no apps had the ability to share content entered into an app with a clinician. For example, sharing safety plans with clinical teams or medical records.

### Privacy

When searching the app store, over half of included apps displayed their privacy policy on the app’s product page (34/52, 65.4%). However, far fewer had their privacy policy integrated into the app itself (19/52, 36.5%), and in some of these, the policy did not load. Nearly a third of apps had no policy available on either the app or product page (15/52, 28.8%). A small number of apps asked users to agree to a set of terms of conditions before using the app, including information about privacy, and limits of the app (9/52, 17.3%). Only two apps allowed users to create a password protected account, however one of these apps required users to pay for an upgrade to use this feature (2/52, 3.8%).

### Design

Many apps included some form of personalization options, with over half prompting users to add text which could be saved and referred back to at later times (30/52, 57.7%). For example, allowing users to enter and save the contact details of support sources. Fewer had the ability for users to upload their own media content, such as photos, music, or voice notes (6/52, 11.5%). Only two (2/52, 3.8%) apps allowed the user to personalize app appearance, for example, choosing background colours. However, one of these required the user to pay for an upgrade, and the other malfunctioned when reviewers attempted to use the feature. Whilst most apps contained only text content, around a quarter contained multimedia content such as videos and audio-recordings (14/52, 26.9%). A few apps offered notifications to the user (11/52, 21.2%), for example, reminders to complete mood monitoring or update safety plans.

### App Content

When assessed against the Essential Features Framework, only two (2/52, 3.8%) apps included content from all domains. Almost three quarters of apps contained four or more domains (37/52, 71.2%). The domain covered by most apps was helpful resources for help-seeking where every app included at least one suggestion of a source of support (52/52, 100.0%), followed by both immediate sources of help-seeking (47/52, 90.4%) and general information about suicide (47/52, 90.4%), positivity and inspiration (33/52, 63.5%), distraction and alternate activities (22/52, 42.3%), safety planning resources (20/52, 38.5%), and wellness content (11/52, 21.2%). The domain covered by the least apps was screening tools (9/52, 17.3%). In the following section app content related to each domain of the Essential Features Framework is discussed in detail. The denominators used within these sections reflect the number of apps covering the domain.

### General Information about Suicide (covered by 47/53, 90.4% of apps)

Most apps included information related to suicide for the user (47/52, 90.4%). Almost all of these apps included information on warning signs that a suicidal crisis may occur (43/47, 91.5%). Nearly half also covered risk factors related to suicide (23/47, 48.9%), with fewer providing information about mental health generally (10/47, 21.3%), or health behaviours such as information about diet, sleep, and exercise (14/47, 29.8%). Over a third of apps contained information related to suicide among marginalized groups (16/47, 34.0%), for example, statistics regarding the risk of attempting suicide among LGBTQIA+ groups. Over three quarters contained information tailored for individuals concerned or supporting someone who may be suicidal (37/47, 78.7%), with a few of these providing information about bereavement through suicide (6/47, 12.8%).

### Wellness (covered by 11/52, 21.2% of apps)

Few apps contained content related to wellness (11/52, 21.2%). Of those, most related to relaxation techniques such as breathing and grounding exercises (10/11, 90.9%), and almost half had mindfulness and meditation resources (5/11, 45.5%). Some of these apps also had space for users to keep a journal or diary (4/11, 36.4%).

### Positivity and Inspiration (covered by 33/52, 63.5% of apps)

Over half of apps included content related to positivity and inspiration (33/52, 63.5%). Most of these included inspirational messaging (30/33, 90.9%) such as encouraging individuals to seek help, reminding users they are not alone, and messages surrounding hope for the future. Some encouraged users to save their reasons for living, usually within a safety plan (13/33, 39.4%). A minority included inspirational quotes from individuals with lived experience (4/33, 12.1%), including individuals who had experienced suicidal thoughts or attempts, or individuals who had supported a person with these experiences.

### Distraction and Alternate Activities (covered by 22/52, 42.3% of apps)

Almost half of apps contained content aimed to distract users from suicidal thoughts, or provide alternate activities for users (22/52, 42.3%). All of these included suggestions of coping strategies, including suggestions for places or activities aimed at distraction (14/22, 63.6%). Most of this information was presented within safety planning tools.

### Safety Planning (covered by 20/52, 38.5% of apps)

Multiple apps allowed users to construct a customizable safety plan (20/52, 38.5%). There was variation in the content of safety plans between apps, however all but one app prompted users to enter the details of trusted contacts in case of crisis (19/20, 95.0%). Some plans requested users to reflect on potential means of suicide method, and how to protect themselves from these when in crisis. However, concerningly some of these apps listed potential means of suicide in this section demonstrating potentially harmful content. Few of the apps containing safety plans allowed the safety plan to be directly shared from the app to another person (6/20, 30.0%).

### Monitoring and Screening Tools (covered by 9/52, 17.3% of apps)

Screening tools were the least common feature among included apps (9/52, 17.3%). Among those containing tools, most involved screening for suicide risk or mental distress (8/9, 88.9%). Examples of screening tools included the Patient Health Questionnaire (PHQ-9) [21], Kessler Psychological Distress Scale (K10) [22], and the Columbia-Suicide Severity Rating Scale (C-SSRS) [23]. Only five tools allowed users to monitor their mood, for example, using a tracker (5/9, 55.6%).

### Helpful Resources (for help-seeking) (covered by 52/52, 100% of apps)

All apps included helpful resources of help-seeking. This included the contact details of suicide prevention phone lines (49/52, 94.2%) and emergency or mental health services (49/52, 94.2%). Many also allowed users to save the contact details of trusted support sources (30/52, 57.7%). This was usually but not always presented within a safety plan. Almost three quarters of apps contained support sources tailored for marginalized groups, or groups who may be at increased risk of suicidal behaviours such as LGBTQIA+ individuals and veterans (38/52, 73.1%). Some apps allowed sources of support to be personalized by user location (11/52, 21.2%), for example, displaying a map with local hospitals and crisis centres.

### Immediate Resources (covered by 47/52, 90.4% of apps)

Most apps gave users access to immediate resources (47/52, 90.4%). Some of these apps had links to emergency support embedded within the app homepage (24/46, 52.2%), most commonly the link to emergency medical support (e.g., 999 in UK, 911 in Canada) or a suicide prevention phone line.

## Discussion

We analyzed the characteristics and content of 52 suicide prevention apps available in the UK and Canada. All apps included at least one type of content recommended for suicide prevention, most commonly sources of support and information about suicide. However, many apps lacked other evidence-based suicide prevention tools such as mood monitoring, screening, wellness tools, and safety planning resources. During the review, the authors experienced multiple issues identifying apps.

Availability of suicide prevention apps were highly transient throughout, with many becoming unavailable during the study process. The instability of healthcare apps has been frequently demonstrated [17, 24, 25]. Specific to suicide prevention apps, in one study, 50% of available suicide prevention apps had changed within 115 days of conducting initial searches [26]. Also, when conducting searches, many apps were far down the app store search results, therefore lacking visibility to potential users. Difficulty finding relevant apps is a longstanding problem for app users [27]. App developers should consider how individuals may search for their app and ensure that an app’s name, description, or category includes explicit mention of terms such as mental health, suicide, safety planning, and other commonly used search terms. This will ensure intended users are able to easily find relevant apps most suitable to their current needs. Without consideration for how users search for and select mobile apps for suicide prevention, app developers risk poor uptake and retention of digital interventions [28].

Similar to previous reviews of suicide prevention apps [15], a significant proportion of apps had not been recently updated or had malfunctioning content. In the context of suicide prevention this may have dangerous implications. Apps targeted at mental health and suicide prevention may benefit from a review by an independent regulator to ensure safe content [15], alongside app developers having a standardized process to ensure app content is regularly updated. A significant number of apps lacked a clear privacy statement. Lack of data privacy, and “clunky” apps have been identified as barriers to use of mental health apps by mental health professionals integrating mobile technology into mental health services [29] and users of such apps [30].

All of the included apps contained at least one type of suicide prevention tool. However, there was a distinct lack in some types of content, for example, the ability for users to access wellness content, complete a safety plan, monitor their mood, or screen for suicide risk (although we emphasize the lack of evidence related to risk prediction of suicide [31]). Additionally, similar to previous reviews of mental health apps, many lacked accessibility [32] or personalization options, which previous research suggests are important facilitators of app use alongside promoting inclusivity in access [33, 34]. Few apps used notifications, which may contribute to poor user retention and engagement. Notifications have been shown to increase user engagement in apps [35, 36], but users of mental health apps have been shown to have mixed opinions as to their appropriateness. For example, notifications may appear as intrusive, reduce the level of privacy due to visibility on a user’s screen, or remind users of the difficulties they are facing [37]. The considerations mentioned above provide evidence for the importance of prioritizing co-production with the intended user group during app development to ensure content meets the preferences and values of users. Important considerations are included in the app evaluation model developed by the American Psychiatric Association. This model reiterates the importance of the accessibility, privacy and security, clinical foundation, usability, and data integration towards therapeutic goals for all mental health apps [38].

No apps identified for this study were designed for integration with mental health services, nor were they connected to any electronic health record system. Embedding apps into clinical practice may present several benefits for service users and staff. This may include the ability for clinicians to collect daily mood monitoring data, patients being able to complete ‘between-session’ activities, and the potential for integration of wearable devices to gather clinical data such as sleep and activity tracking [39]. A scoping review has examined the literature on information and communication technology-based suicide prevention interventions (including apps) that have been implemented in clinical settings [40]. The reviewers excluded apps available in app stores if they were not implemented in clinical settings and identified ten suicide prevention apps which were implemented in multiple clinical settings (e.g., inpatient, outpatient, and community), and groups of patients (e.g., adults, children and young adults). Several barriers were identified as hindering integration such as lack of skills to use technology, unstable internet connection, and lack of buy-in. These challenges are not unique to technologies for suicide prevention or mental health; many other mobile health interventions face challenges with adoption and can be abandoned soon after initiating use [41]. As such, the significance of implementation efforts cannot be underestimated. These efforts involve analyzing the barriers and facilitators for the successful integration of suicide prevention app-based interventions into routine clinical practice and developing targeted approaches to address these barriers while leveraging facilitators. [40].

## Conclusions

This article has analyzed the content and features of suicide prevention apps available in UK and Canada. Although all apps contained some form of suicide prevention tools, many still did not contain evidence-based features. Furthermore, suicide prevention apps continue to lack personalization and accessibility options. The findings suggest implications for future evidence-based suicide prevention app development. There is the potential for apps to be utilized as an adjunct to clinical care, however a cautious approach must be taken if apps link to clinical notes and/or electronic health records, and apps should not be considered as an alternative to provision of therapeutic interventions with appropriately trained staff. The development of future suicide preventions apps provides an opportunity for authentic co-production with both service users and staff, and a novel way to evaluate data in real time.

## Data Availability

Two appendices are available as supplementary files which include the data extraction template used for the app store review, a list of included apps, apps excluded at full review stage, and reasons for exclusion. Additional data is available upon request to the corresponding author.

## Author Contributions

This study was conceptualized by KL and GS. App searching and screening was then conducted by LBP, SG, JK, and DS. Data extraction was completed by LBP, SG, and JK followed by data analysis by LBP and SG. The original draft of this paper was written by LBP, SG, JK, and DS and was reviewed and edited by KL, GS, and LS. All authors read and agreed upon the final version of the article.

## Funding

This research received support from the Centre for Addiction and Mental Health (CAMH) and Oxford Health NHS Foundation Trust (OHFT).

### Abbreviations

LGBTQIA+: lesbian, gay, bisexual, transgender, queer, intersex, asexual, and other identities
UK: United Kingdom
PRISMA: preferred reporting items for systematic reviews and meta-analyses
PHQ-9: patient health questionnaire
K10: Kessler Psychological Distress Scale
C-SSRS: Columbia-suicide severity rating scale

## Data Sharing Statement

**Supplementary File 1** See attached.

**Supplementary File 2** See attached.

