## Supplemental File 1 for "Characteristics of Suicide Prevention Apps: A Content Analysis of Apps Available in Canada and the United Kingdom"

**Supplementary File 1: Template Data Extraction Form**

| Category | Extraction Point | Results |
|---|---|---|
| General Information | Name of App | |
| | Name of Reviewer and App Format (i.e., Apple or Android) | |
| | Reviewer and App Format for Verification (i.e., Apple or Android) | |
| | Did the App Become Unavailable During the Screening Process? | |
| | Number of Downloads | |
| | Date Last Updated | |
| | Average User Reviews (out of 5) | |
| | Developer Country | |
| | App Category (e.g., Health & Fitness, Reference, etc.) | |
| | Suggested Age Range | |
| | Can User Create an Account (e.g., Login Details) | |
| | Ability to Save Content in the App | |
| | Can App Content be Password Protected? | |
| | Who is the Target User? | |
| | Are There App Instructions for Users? | |
| | Reviewer Experienced Errors When Trying Out App | |
| | Content Available in Additional Languages to English | |
| FRAMEWORK DOMAIN 1: General Information about Suicide | Risk Factors for Suicide | |
| | Warning Signs | |
| | General Information About Suicide | |
| | Myths About Suicide | |
| | General Information About Mental Health | |
| | Information About Health Behaviours | |
| | Information About Marginalised Groups | |
| | Information for Carers/People Worried About Someone Else | |
| | Information on Life After Attempt/Recovery | |
| | Information About Bereavement | |
| | Resources Related to COVID-19 | |
| | Summary (Select If Any of the Sections Has a “Yes”) | |

| | |
|---|---|
| **FRAMEWORK DOMAIN 2: Safety Planning** | Customisable Safety Plan |
| | Pre-made Safety Plan (Without Customisation) |
| | Can the Users' Contacts be Linked to Safety Plan? |
| | Can Safety Plan be Shared Using App? |
| | Summary (Select If Any of the Sections Has a "Yes") |
| **FRAMEWORK DOMAIN 3: Positivity and Inspiration** | Quotes from Individuals With Lived Experience |
| | Reasons for Living |
| | Positive Messaging |
| | Summary (Select If Any of the Sections Has a "Yes") |
| **FRAMEWORK DOMAIN 4: Helpful Resources (for Help-seeking)** | Information on Suicide Prevention Helplines |
| | Information on Emergency Departments/Crisis Centres |
| | Information on General Mental Health Organisations/Helplines |
| | Information on Services Available for Marginalised Groups |
| | Organisations Can Be Personalised Using Location of User |
| | User Can Add Own Support Contacts |
| | Summary (Select If Any of the Sections Has a "Yes") |
| **FRAMEWORK DOMAIN 5: Immediate Help-seeking** | Click to Access Emergency Support (Select If Either of the Sections Has a "Yes") |
| | Single Click to Emergency Support on Home Screen |
| **FRAMEWORK DOMAIN 6: Wellness** | Mindfulness Techniques/Meditation |
| | Relaxation Techniques |
| | Information About Values and Setting Goals Towards These |
| | Journaling (NOT Including Mood Monitoring) |
| | Summary (Select If Any of the Sections Has a "Yes") |

| | |
|---|---|
| **FRAMEWORK DOMAIN 7: Distraction & Alternate Activities** | Coping Strategies |
| | Distraction Techniques or Activities |
| | Summary (Select If Any of the Sections Has a “Yes”) |
| **FRAMEWORK DOMAIN 8: Screening Tools** | Mood Monitoring |
| | Tracked Mood Monitoring (e.g., Provides Reports Over Time) |
| | Screening for Mental Health Conditions or Suicide Risk or Suicidal Thoughts/Behaviours |
| | Summary (Select If Any of the Sections Has a “Yes”) |
| **Extraction Points from Service User and Staff Consultation** | Can the App Be Linked to Clinician/Medical Records? |
| | Is There Content for Carers/People Concerned? |
| | Are There Accessibility Options? |
| | Is App Content Shown in Multiple Formats (e.g., Text/Videos/Audio)? |
| | Can Users’ Own Resources Be Added (e.g, Pictures, Audio)? |
| | Does the App State They Are Collecting Data on User? (Reviewer to Search in Privacy Policy if One is Given) |
| | Does App Ask for Consent (e.g., Agree to Terms and Conditions)? |
| | Privacy/Data Policy in App Store |
| | Privacy/Data Policy in App |
| | Does App Provide Notifications? |
| | Can the Appearance of the App Be Personalised? |
