## Supplemental File 2 for "Characteristics of Suicide Prevention Apps: A Content Analysis of Apps Available in Canada and the United Kingdom"

**Supplementary File 2: List of Included Apps**

1. A Friend Asks
2. Alaska Careline
3. Albany County Hope
4. Am I? My Safety Plan
5. Anemone Crisis App
6. Be Safe
7. Be Safe by mindyourmind
8. Calm Care
9. Case Western Reserve Reach Out
10. Colombia Protocol
11. Embracing Life
12. First Step Oregon
13. HELP App-Prevention Resources
14. Hope by CAMH
15. iHelp Sunshine Coast
16. Is S/O Suicidal?
17. Lakeland CC Reach Out
18. MoodTools - Depression Aid
19. MS DMH: Shatter the Silence
20. My Tools - suicide.ca
21. National Louis Univ - Reach Out
22. Operation Life
23. Operation Reach Out
24. Prevent Suicide - D&G
25. Prevent Suicide - Highland
26. Prevent Suicide- NE Scotland
27. R U Suicidal?
28. Relief Link
29. ReMinder Suicide Safety Plan
30. SecurApp
31. Sinclair College Hope Link
32. SP Lanarkshire
33. Stanley-Brown Safety Plan
34. Stay Alive
35. Suicide Prevention App
36. Suicide Safety Plan
37. Suicide? Help! Tayside
38. The Fire Watch
39. Tri-C Help Is Here
40. TUFMinds
41. Univ of Cincinnati Reach Out
42. Upenyu: Mental Health App
43. We Care 59th Ordnance Brigade
44. We Care CASCOM
45. We Care Fort Campbell
46. We Care Fort Jackson

47. We Care Fort Rucker (Renamed as We Care Fort Novosel during study period)
48. We Care, Ft Bragg
49. We Care JBM-HH
50. We Care JBSA
51. We Care USACE
52. Yellow Ribbon App

**Apps Excluded at Full Review Stage and Reasons for Exclusion**

| App Name | Reason for Exclusion |
|---|---|
| Be Calm | Cannot log in to app for access, errors prevent extraction |
| Brighter Side | Cannot log in to app for access, errors prevent extraction |
| Dr.Mind: Mental Health Tests | Cannot log in to app for access, errors prevent extraction |
| INSIST | Cannot log in to app for access, errors prevent extraction |
| My Companion Journal | Cannot log in to app for access, errors prevent extraction |
| My SP | Cannot log in to app for access, errors prevent extraction |
| PsychStar | Cannot log in to app for access, errors prevent extraction |
| The Lifeline | Cannot log in to app for access, errors prevent extraction |
| distrACT | Focus is not on suicide prevention |
| My Mental Health Crisis Plan | Focus is not on suicide prevention |
| DMHS: Interactive Suicide Prevention | No longer available |
| A Teen Suicide Prevention Anim | No longer available |
| AlachuaTalk | No longer available |
| Better Stop Suicide | No longer available |
| Centar Srce | No longer available |
| DMHS: Suicide Prevention and Crisis | No longer available |
| Hello Lifeline | No longer available |
| My Mental Health Risk | No longer available |
| PMCS Combating Suicide | No longer available |
| Psychiatry Pro-Diagnosis,Info,Treatment,CBT & DBT | No longer available |
| Safety Net | No longer available |

| | |
|---|---|
| SafetyNet: Your Suicide Prev | No longer available |
| Suicide Lifeguard | No longer available |
| Suicide Prevention -Ways to Heal | No longer available |
| Min Livlina | Not in English language |

**Reasons for Exclusion Counts:**

- No longer available N=14
- Cannot log in to app for access, errors prevent extraction= N=8
- Focus is not on suicide prevention N=2
- Not in English language N=1
